# Temporal Geospatial Analysis of COVID-19 Pre-infection Determinants of Risk in South Carolina

**DOI:** 10.1101/2021.08.02.21261500

**Authors:** Tianchu Lyu, Nicole Hair, Nicholas Yell, Zhenlong Li, Shan Qiao, Chen Liang, Xiaoming Li

**Author notes:** Equal contribution (Tianchu Lyu, Nicole Hair).

## Abstract

**Introduction:** Disparities and their geospatial patterns exist in coronavirus disease 2019 (COVID-19) morbidity and mortality for people who are engaged with clinical care. However, studies centered on viral infection cases are scarce. It remains unclear with respect to the disparity structure, its geospatial characteristics, and the pre-infection determinants of risk (PIDRs) for people with the infection. This work aimed to assess the geospatial associations between PIDRs and COVID-19 infection at the county level in South Carolina by different timepoints during the pandemic.

**Method:** We used global models including spatial error model (SEM), spatial lag model (SLM), and conditional autoregressive model (CAR), as well as geographically weighted regression model (GWR) as a local model to examine the associations between COVID-19 infection rate and PIDRs. The data were retrieved from multiple sources including USAFacts, US Census Bureau, and Population Estimates Program.

**Results:** The percentage of males and the percentage of the unemployed population were statistically significant (p values < 0.05) with positive coefficients in the three global models (SEM, SLM, CAR) throughout the time. The percentage of white population and obesity rate showed divergent spatial correlations at different times of the pandemic. GWR models consistently have a better model fit than global models, suggesting non-stationary correlations between a region and its neighbors.

**Conclusion:** Characterized by temporal-geospatial patterns, disparities and their PIDRs exist in COVID-19 incidence at the county level in South Carolina. The temporal-geospatial structure of disparities and their PIDRs found in COVID-19 incidence are different from mortality and morbidity for patients who are connected with clinical care. Our findings provided important evidence for prioritizing different populations and developing tailored interventions at different times of the pandemic. These findings provided implications on containing early viral transmission and mitigating consequences of infectious disease outbreaks for possible future pandemics.

## Introduction

Coronavirus disease 2019 (COVID-19), caused by the Severe Acute Respiratory Syndrome Coronavirus 2 (SARS-CoV-2), is a highly contagious disease that has caused widespread panic and concern across the globe. COVID-19 was the third leading cause of death in 2020. The death rate increased by 15.9% from 2019 to 2020.^1^ As of June, 2020, there have been 34.1 million confirmed cases and 609 thousand deaths due to the COVID-19 in the US.^1-3^ Additionally, COVID-19 has a profound impact on social life and the economy, as closing businesses and social distancing have been a common practice to stopping the spread of the disease. The US real GDP decreased by 3.5% in 2020 and was projected to lose at least $3.2 trillion due to COVID-19 in a two-year course.^4,5^

The burdens of COVID-19 have not been borne equally. Some populations face increased risk for COVID-19 morbidity and mortality. A sizable collection of studies have reported disparities in the clinical outcomes of patients with COVID-19. For example, studies using inpatient data found severe disease progression and poor clinical outcomes of COVID-19 patients to be associated with a set of underlying medical conditions (e.g., hypertension, diabetes, asthma, and heart, liver, and respiratory illnesses), demographics (e.g., male, older age, race/ethnic minority), and social determinants of health (SDOHs) (e.g., lower education and income).^6-10^ A large cohort in Louisiana comprised of 3,481 COVID-19 patients reported that 76.9% of the hospitalized cases and 70.6% of the death cases were black, whereas only 31% of the population are black people.^11^ While these studies have provided a critical evidence base of disparities in COVID-19 clinical outcomes and implications on medical care for addressing the disparities, they provided limited implications for disparities in the risk of exposure to COVID-19 for the following reasons. First, the findings of these studies are applicable for hospitalized patients but may not be generalizable for outpatients, individuals with mild symptoms, and asymptomatic individuals since they are based on the secondary analysis of inpatient data. The omission in outpatients and individuals with laboratory-confirmed COVID-19 infections but have no clinic visits will harm the potential opportunity of exploring risk factors for these populations.^12^ Second, using disease severity as the outcome variable does not provide implications on SARS-CoV-2 infection and transmission. For example, SARS-CoV-2 transmits more easily in regions with a large proportion of the younger population, yet the elderly were found at a higher risk of developing poor clinical outcomes.^13^

Therefore, it is equivalently important to curate an evidence base for disparities in the risk of exposure to COVID-19 and the pre-infection determinants of risk (PIDRs) (e.g., demographics, socioeconomic, and prevalence of diseases related to COVID-19 infection).^14-19^ Such an evidence base can be used for understanding disease transmission patterns, identifying vulnerable populations, and proactively mitigating disparities in future pandemics.^20^ Existing studies have reported demographic and socioeconomic factors to be related to disparities in the risk of exposure to COVID-19. Different combinations of those determinants lead to different health attributes (e.g., health behaviors and physical conditions), thus influence the spread of the virus. For example, high-deprivation areas have higher rates of hospitalization and testing.^16^ People with higher income are more likely to engage in self-protecting behavior during the COVID-19 pandemic.^17^ Another study reported that the behaviors of wearing masks and using hand hygiene are associated with female gender and higher education level among students in Chinese population.^18^ In a primary care cohort, researchers observed a higher risk of COVID-19 infection among people aged 40-64 years, in the male gender, black race, and living in urban areas.^14^ Incorporating census tract level data with the COVID-19 dataset, Hawkins and colleagues examined the association between socioeconomic status variables and COVID-19 cases at the county level across the US and found a lower education level and higher percentage of black residents to be risk factors for the infection.^15^

To further explore the associations between these PIDRs and COVID-19 transmission, geospatial information is needed as geographic differences exist across states, counties, and communities in the timing of SARS-CoV-2 introduction, which are further characterized by population density, local policies, and population composition.^13^ Particularly, understanding PIDRs and their geospatial epidemiology is urgently needed for rural states such as South Carolina as the state has disproportionally low healthcare capacity and a high disease burden. It may also provide timely information for post-COVID-19 care, given the emerging reports on the heterogeneity of symptoms in individuals with post-acute sequelae of SARS-CoV-2 infection (PASC).^21,22^ Although the spatially dynamic nature of infectious diseases (e.g., different spatial patterns of transmission) makes geospatial analysis a valuable tool to unveil the epidemiology,^23-26^ there have been limited studies reporting the geospatial characteristics of PIDRs.^13,27-30^ Several studies have reported ethnic minorities, age, and other social vulnerabilities to be associated with a higher COVID-19 infection, yet spatial patterns were generally not included in the statistical models as independent variables. ^27,29,30^ Fortaleza and colleagues used multivariate regression and found that demographic density and distance from the state capital are robust predictors of COVID-19 prevalence in Brazil.^28^ Another study built a correlation matrix between socioeconomic determinants and COVID-19 case rate across the US and found population density to be highly correlated with COVID-19 prevalence.^13^

Although these studies have collectively suggested possible geospatial characteristics among the disparities in virus transmissions, spatial autocorrelation is generally not included in the statistical models, which limits the statistical power of the findings. The spatial autocorrelation, including global modeling and local modeling approaches, enables the correlation measure of a variable (e.g., PIDRs) with itself across different regions. Spatial global models assume a stationary correlation between a region and its neighbors, whereas spatial local models assume non-stationary correlations between a region and different neighbors. Among a few preliminary studies that adopted spatial autocorrelation, Mollalo and colleagues examined the association between COVID-19 incidence rate and four county-level explanatory determinants including income inequality, median household income, the percentage of nurse practitioners, and the percentage of the black female population to the total female population across the US.^31^ The authors started out with a set of 35 socioeconomic, behavioral, topographic, and demographic explanatory variables. After a stepwise forward procedure and correlation analysis, they choose to keep four of these variables in their final model and found that geographically weighted regression (GWR) models best explained the variations, suggesting the existence of spatial autocorrelation and different vulnerabilities across the counties. Despite highly appropriate methods, the study could have better interpreted the disparity structure if demographic determinants such as age, gender, and race were included in the analysis. Additionally, because these studies are based on analyses of panel data, they did not specify whether and how disparities and the PIDRs vary by different times as the pandemic evolves. Moreover, there is increased endogeneity in these analyses because they focused on large geographic regions by which different regional policies might have a greater impact on the COVID-19 prevalence as compared with the explanatory variables, as existing evidence suggests that government responses and socioeconomic determinants have played an important role in the transmission of SARS-Cov-2, which differs geographically.^32^ Another similar study instead included demographics in PIDRs but still suffered from the same endogeneity problem.^33^

Building on these existing studies, we sought to assess the association between PIDRs (including demographics, socioeconomic, and prevalence of diseases related to COVID-19 infection) and COVID-19 infection at the county level in South Carolina by different timepoints amid the pandemic. The heterogeneity in the virus spread in South Carolina suggests that different PIDRs in certain areas could enhance or inhibit the transmission of COVID-19. Within the smaller geographic scale of one state, the heterogeneous impacts of different regional policies could be largely mitigated, and the data abundance is sufficient for conducting geospatial analyses. Anticipated findings of this study would inform an evidence base for temporal geospatial disparities in the risk of exposure to COVID-19 and the associated PIDRs. The identified PIDRs may also shed light on the populations and regions vulnerable to PASC in South Carolina during post-COVID-19 care.

## Methods

### Model selection

We selected six time windows to represent the COVID-19 cases at different times of the pandemic. South Carolina began tracking COVID-19 cases in early March of 2020 and the number of daily new cases began to rise until July 2020 when the daily number of new cases began to fluctuate. We calculated the average cumulated case numbers in a sliding window of seven days (15^th^ ± 3 days) for every month between July and December of 2020.

The US Centers for Disease Control and Prevention (CDC) has provided a list of risk factors of COVID-19 severity such as age and existing medical conditions.^34^ As discussed in the Introduction, the PIDRs for COVID-19 severity can be very different from the PIDR for COVID-19 infection. Based on previous studies, Snyder and Parks presented a well-developed risk factor index framework for COVID-19 community vulnerability which was defined as “the potential decrease in the wellbeing of a community before and during/after the pandemic, taking into account health, social, and economic conditions”.^35-37^ The index is divided into four major sections (e.g., ecological, social, health, and economic).^36^ Inspired by their study and based on data availability in South Carolina, we started off with 15 different variables related to the four sections of the index including sex, age, race, median household income, population density, uninsured rate, poverty percentage, high school degree rate, college degree rate, unemployment rate, physical inactivity rate, obesity rate, smoking prevalence, medical doctors per 10,000 people, and nurse practitioner per 10,000 people (Table 1). Among the candidate variables, age, gender, and population density represent ecological variables; uninsured rate, education levels, race, medical doctor abundance, and nurse practitioner abundance are social variables; obesity rate, physical inactivity rate, and smoking prevalence are health variables; and income, poverty rate, and unemployment rate are economic variables.

**Table 1.**
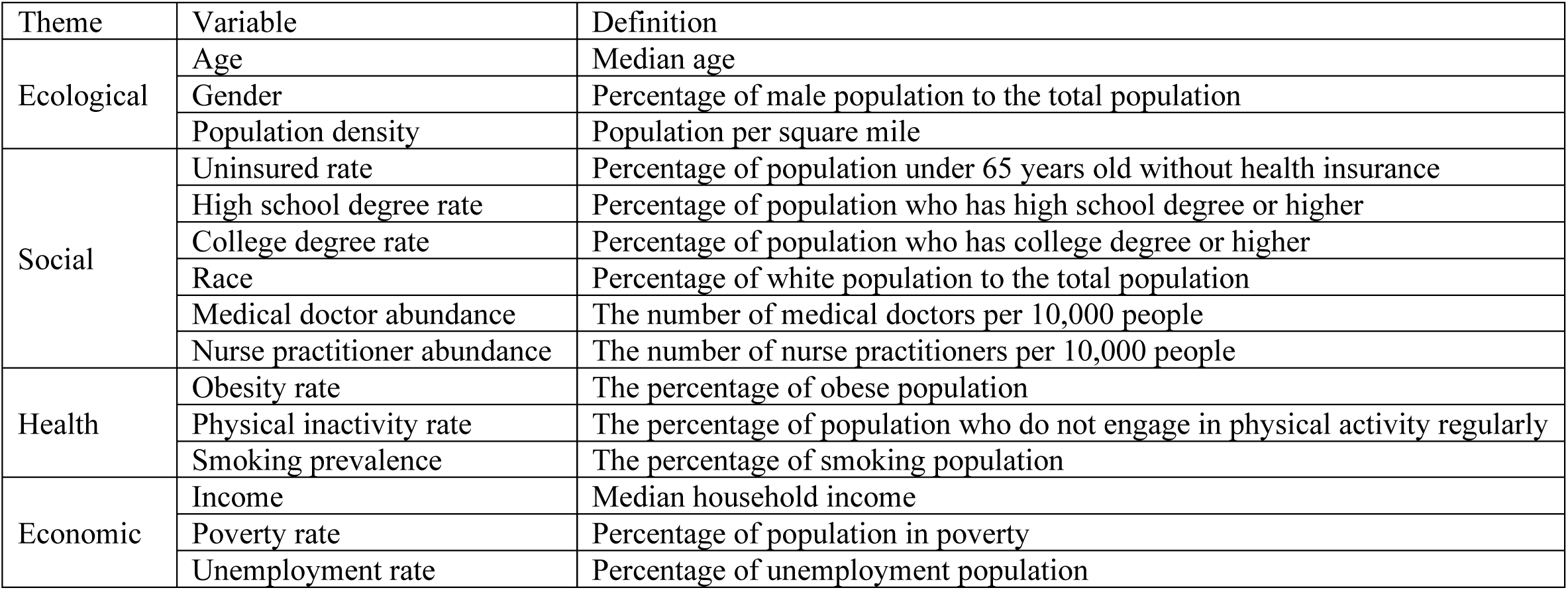
Candidate explanatory variables and definitions

| Theme | Variable | Definition |
| --- | --- | --- |
| Ecological | Age | Median age |
|  | Gender | Percentage of male population to the total population |
|  | Population density | Population per square mile |
| Social | Uninsured rate | Percentage of population under 65 years old without health insurance |
|  | High school degree rate | Percentage of population who has high school degree or higher |
|  | College degree rate | Percentage of population who has college degree or higher |
|  | Race | Percentage of white population to the total population |
|  | Medical doctor abundance | The number of medical doctors per 10,000 people |
|  | Nurse practitioner abundance | The number of nurse practitioners per 10,000 people |
| Health | Obesity rate | The percentage of obese population |
|  | Physical inactivity rate | The percentage of population who do not engage in physical activity regularly |
|  | Smoking prevalence | The percentage of smoking population |
| Economic | Income | Median household income |
|  | Poverty rate | Percentage of population in poverty |
|  | Unemployment rate | Percentage of unemployment population |

We then tested multicollinearity across the candidate variables and finetuned the final model with demographic variables including age, gender, race, and socioeconomic variables including unemployment rate, uninsured rate, college degree rate, obesity rate, and nurse practitioner per 10,000 people. Specifically, we excluded only the variables (i.e., median household income, population density, poverty percentage, high school degree rate, physical inactivity rate, smoking prevalence, medical doctors per 10,000 people) that were highly correlated with other variables (correlation coefficient > 0.7) in this step. We set the criterion not very strict for two reasons: (1) Because we used spatial regression afterward, multicollinearity would be different after we incorporated spatial autocorrelation. (2) We wanted to include as many variables as possible to better represent the Snyder and Parks’s index, so the results could be more intuitive and interpretable.

### Data sources

The data sources used in this work varied. The age variable was extracted from US Census Population and Housing Unit Estimates, 2010-2018. Gender, race, and college degree rate were extracted from US Census Bureau, Population Estimates Program (PEP), and American Community Survey (ACS), updated July 1, 2019. The unemployment rate and poverty rate were retrieved from US Census Bureau, Small Area Income and Poverty Estimates (SAIPE) Program (2019). The uninsured rate was retrieved from US Census Bureau, Small Area Health Insurance Estimates (SAHIE) Program (2018). The obesity rate was retrieved from US CDC Diabetes County Data Indicators, 2006-2017. The nurse practitioner number was retrieved from Health Resources & Services Administration (HRSA) Area Health Resources Files, 2017 and 2018. The confirmed cases number of COVID-19 from July 2020 to December 2020 was obtained USAFacts, which is also the data source that US CDC use. Log transformation was applied for the dependent variable and the explanatory variables to normalize data.

### Spatial regression models

We calculated spatial weights using queen contiguity which defines neighbors by the presence of shared edges and vertices. Figure 1 shows the county map of South Carolina with the links between each neighbor (i.e., county). Spatial modeling was used to describe the relationship between the COVID-19 cases and factors since we were able to split the data into counties and wanted to focus on how and why the cases differed across South Carolina. The following spatial models were used to fit our data: spatial error model (SEM), spatial lag model (SLM), conditional autoregressive (CAR) model, and GWR model. We used SEM to observe spatial autocorrelation between the residuals of neighboring counties, which incorporates spatial effects through the error term. SLM applies spatial dependence by adding a spatially lagged response variable as an additional predictor on the linear model equation. This model assumes that the COVID-19 incidence rate in one county is directly influenced by the COVID-19 incidence rates in its neighboring counties. If positive spatial lag is observed in SLM, it would suggest that COVID-19 incidence rates in neighboring counties covary. CAR model relies on the conditional distribution of the spatial error terms and assumes the region is a function of its neighbors but not the neighbors of neighbors (i.e., first-order dependency). We used the GWR method to examine the local models, which is based on kernel-weighted regression and allows for parameters to vary spatially.^38^

**Figure 1.**
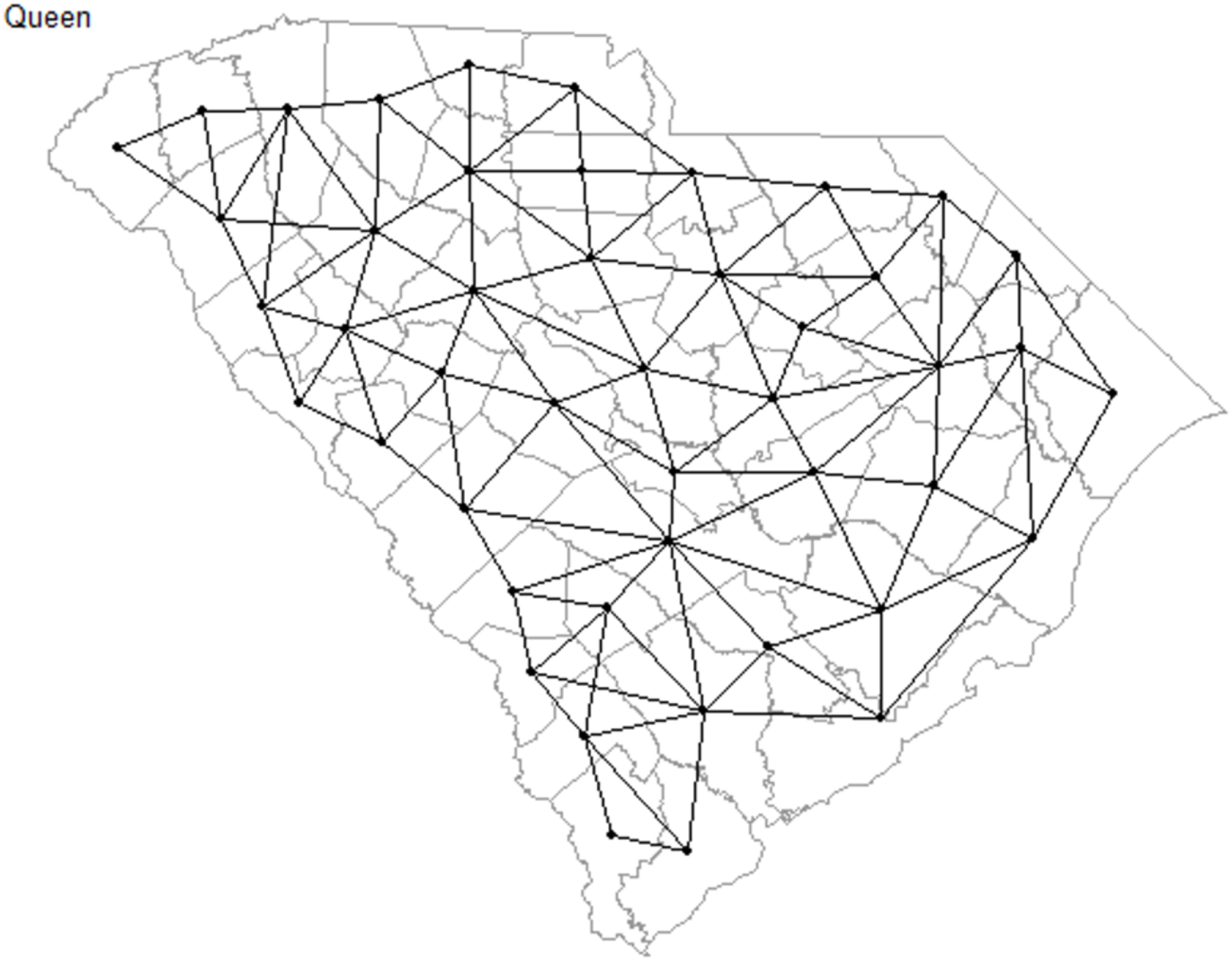
Contiguity Queen Neighbors of Counties in South Carolina.

## Results

### Distribution of COVID-19 cases and covariates

After model selection, the final model contained eight explanatory variables, namely male percentage, percentage of the white population, median age, college degree rate, obesity rate, unemployment rate, uninsured rate, and nursing practitioner abundance. We summarized and showed maps of distributions for all the variables in the model (Figure 2). To make the descriptive map comparison between variables easier, we held back the temporal dimension and used average COVID-19 incidences per 1,000 people around July 15^th^. In Figure 2, we could observe certain levels of similarities between COVID-19 cases and different independent variables including demographic and socioeconomic variables. For example, the map of gender and age were congruent with the map of COVID-19 incidence rate. The map of the obesity rate showed a nearly opposite pattern compared to the map of COVID-19 cases.

**Figure 2.**
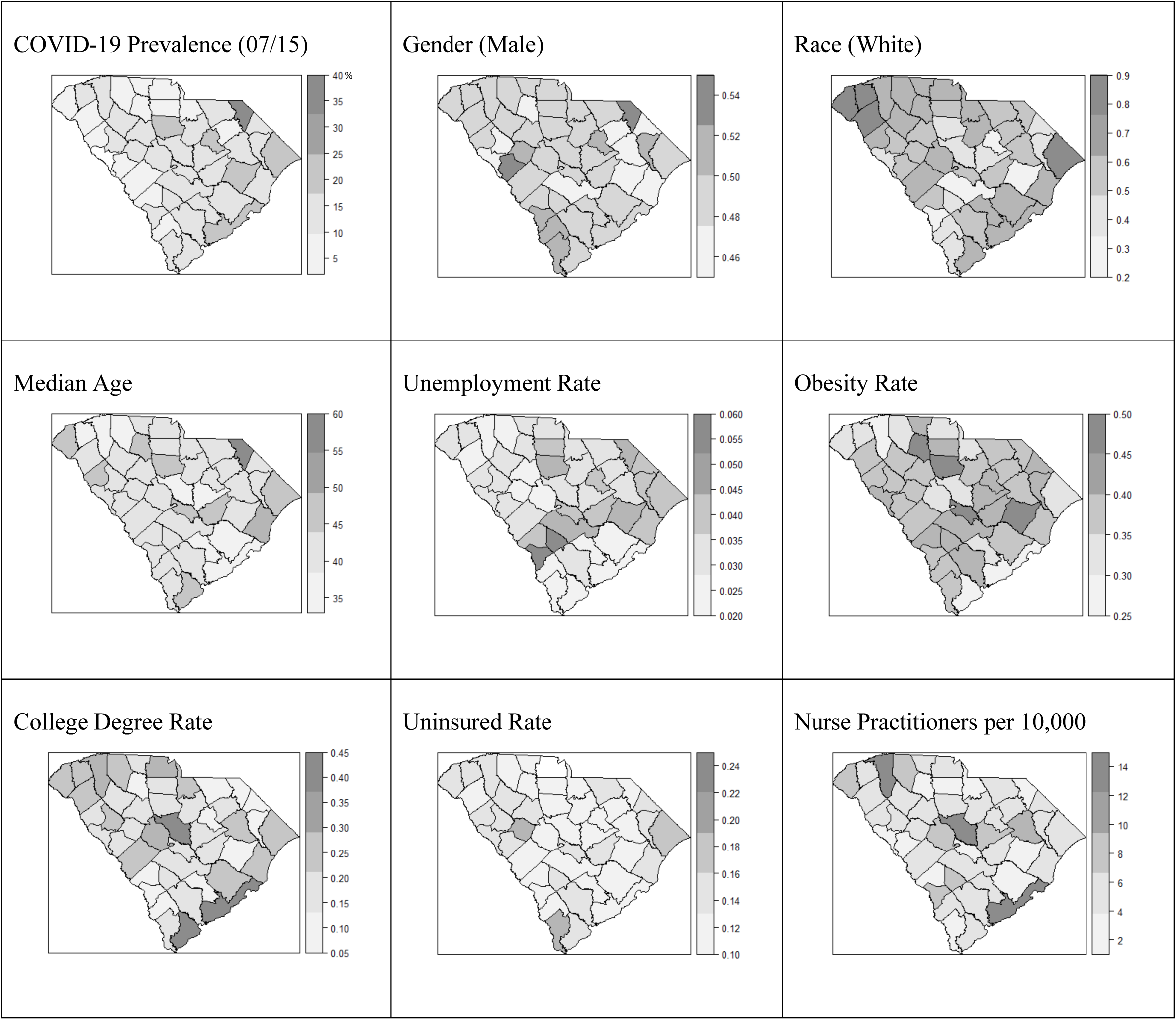
Distribution maps of the variables in the model.

### Global models for spatial correlation

When considering the temporal dimension, four geospatial models were built to examine the spatial correlation of COVID-19 incidence across the counties in South Carolina including SEM, SLM, CAR model, and GWR model. The significant results from Moran’s I test (p values < 0.05) suggested the existence of spatial autocorrelation. We summarized the coefficients of the variables and corresponding p values for global models in Tables 2, 3, and 4 (i.e., SEM, SLM, and CAR model). All the models were significant at 0.05 level generally, indicating that spatial autocorrelations did show within the error terms. The percentage of males and the percentage of the unemployed population were statistically significant (p values < 0.05) with positive coefficients in the three global models throughout the time, while other variables were not (Table 2, 3, and 4). Interestingly, the spatial correlations between COVID-19 cases and the percentage of white population and obesity, respectively, flipped over the course of the pandemic. The percentage of the white population started negatively correlated with COVID-19 cases in July (SLM) and/or August (SEM, CAR) but became positively correlated with COVID-19 cases in September (CAR) and October (SEM, SLM) through December. The percentage of obesity started negatively correlated with COVID-19 cases as early as July and became positively correlated October through December in SEM and CAR, yet this pattern did not show in SLM.

**Table 2.** Coefficients of the explanatory variables in SEM

|  | July 15 |  | August 15 |  | September 15 |  | October 15 |  | November 15 |  | December 15 |  |
| --- | --- | --- | --- | --- | --- | --- | --- | --- | --- | --- | --- | --- |
|  | Coeff. | p value | Coeff. | p value | Coeff. | p value | Coeff. | p value | Coeff. | p value | Coeff. | p value |
| **Male percentage** | 9.26 | *0.014 | 7.72 | *0.013 | 9.65 | **0.002 | 11.30 | **0.000 | 11.92 | **0.000 | 11.84 | **0.000 |
| **White percentage** | 0.21 | 0.970 | -0.06 | 0.910 | -0.05 | 0.966 | 0.35 | 0.407 | 0.73 | 0.069 | 1.17 | **0.003 |
| **Median age** | -0.08 | 0.910 | 0.16 | 0.793 | 0.03 | 0.960 | 0.09 | 0.855 | -0.03 | 0.952 | -0.19 | 0.694 |
| **College degree rate** | 0.53 | 0.273 | 0.39 | 0.313 | 0.45 | 0.228 | 0.35 | 0.263 | 0.34 | 0.243 | 0.38 | 0.198 |
| **Obesity rate** | -2.17 | 0.198 | -0.76 | 0.589 | -0.25 | 0.846 | 0.32 | 0.765 | 0.58 | 0.568 | 0.68 | 0.467 |
| **Unemployment rate** | 2.88 | **0.003 | 2.07 | **0.009 | 2.19 | **0.004 | 2.05 | **0.001 | 2.10 | **0.000 | 2.28 | **0.000 |
| **Uninsured rate** | 1.91 | 0.052 | 0.49 | 0.539 | 0.12 | 0.875 | -0.25 | 0.689 | -0.46 | 0.438 | -0.67 | 0.260 |
| **NP abundance** | 0.08 | 0.575 | 0.04 | 0.737 | 0.07 | 0.490 | 0.12 | 0.160 | 0.13 | 0.111 | 0.13 | 0.106 |

NP: nurse practitioner
\*: p < 0.05
\*\*: p < 0.01

**Table 3.** Coefficients of the explanatory variables in SLM

|  | July 15 |  | August 15 |  | September 15 |  | October 15 |  | November 15 |  | December 15 |  |
| --- | --- | --- | --- | --- | --- | --- | --- | --- | --- | --- | --- | --- |
|  | Coeff. | p value | Coeff. | p value | Coeff. | p value | Coeff. | p value | Coeff. | p value | Coeff. | p value |
| **Male percentage** | 10.70 | *0.015 | 9.28 | **0.008 | 9.65 | **0.002 | 12.81 | **0.000 | 13.30 | **0.000 | 13.16 | **0.000 |
| **White percentage** | -0.07 | 0.911 | -0.48 | 0.348 | -0.02 | 0.966 | 0.18 | 0.646 | 0.66 | 0.077 | 1.21 | **0.001 |
| **Median age** | 0.04 | 0.962 | 0.41 | 0.537 | 0.03 | 0.960 | 0.26 | 0.604 | 0.11 | 0.815 | -0.10 | 0.840 |
| **College degree rate** | 0.44 | 0.372 | 0.22 | 0.583 | 0.45 | 0.228 | 0.15 | 0.608 | 0.14 | 0.611 | 0.19 | 0.522 |
| **Obesity rate** | -3.16 | 0.076 | -2.09 | 0.143 | -0.25 | 0.846 | -0.86 | 0.431 | -0.52 | 0.618 | -0.16 | 0.882 |
| **Unemployment rate** | 2.83 | **0.003 | 1.93 | *0.011 | 2.19 | **0.004 | 2.01 | **0.001 | 2.09 | **0.000 | 2.31 | **0.000 |
| **Uninsured rate** | 1.75 | 0.081 | 0.64 | 0.419 | 0.12 | 0.875 | 0.09 | 0.882 | -0.14 | 0.815 | -0.37 | 0.528 |
| **NP abundance** | 0.07 | 0.640 | 0.03 | 0.801 | 0.07 | 0.490 | 0.11 | 0.213 | 0.12 | 0.149 | 0.13 | 0.124 |

NP: nurse practitioner
\*: p < 0.05
\*\*: p < 0.01

**Table 4.** Coefficients of the explanatory variables in CAR model

|  | July 15 |  | August 15 |  | September 15 |  | October 15 |  | November 15 |  | December 15 |  |
| --- | --- | --- | --- | --- | --- | --- | --- | --- | --- | --- | --- | --- |
|  | Coeff. | p value | Coeff. | p value | Coeff. | p value | Coeff. | p value | Coeff. | p value | Coeff. | p value |
| **Male percentage** | 9.59 | *0.012 | 8.10 | **0.009 | 9.97 | **0.001 | 11.39 | **0.000 | 12.01 | **0.000 | 11.97 | **0.000 |
| **White percentage** | 0.08 | 0.901 | -0.10 | 0.849 | 0.01 | 0.988 | 0.33 | 0.445 | 0.71 | 0.079 | 1.18 | **0.003 |
| **Median age** | -0.04 | 0.953 | 0.23 | 0.700 | 0.07 | 0.908 | 0.12 | 0.813 | 0.00 | 0.996 | -0.17 | 0.727 |
| **College degree rate** | 0.32 | 0.500 | 0.00 | 0.568 | 0.30 | 0.422 | 0.33 | 0.299 | 0.32 | 0.275 | 0.35 | 0.228 |
| **Obesity rate** | -3.13 | 0.065 | -1.61 | 0.242 | -1.09 | 0.402 | 0.24 | 0.821 | 0.53 | 0.598 | 0.66 | 0.512 |
| **Unemployment rate** | 2.82 | **0.002 | 1.99 | **0.008 | 2.17 | **0.003 | 2.02 | **0.001 | 2.07 | **0.000 | 2.27 | **0.000 |
| **Uninsured rate** | 2.16 | *0.026 | 0.79 | 0.317 | 0.43 | 0.568 | -0.22 | 0.728 | -0.44 | 0.461 | -0.64 | 0.279 |
| **NP abundance** | 0.08 | 0.565 | 0.03 | 0.784 | 0.05 | 0.613 | 0.13 | 0.146 | 0.13 | 0.096 | 0.14 | 0.091 |

NP: nurse practitioner
\*: $p < 0.05$
\*\* : $p < 0.01$

### Local models for spatial correlation

The results of the GWR model were summarized in Table 5. In the GWR models, the calculated bandwidths were 60.87 km for July 15 and 154.41 km for the rest time points. Taking July 15 as an example, the GWR model possessed the lowest Akaike Information Criterion (AIC) value of 51.24, compared to the global models such as SEM (AIC = 61.59), SLM (AIC = 58.72), and CAR model (AIC = 59.39) (Table 6). A smaller AIC indicates a better model fitting when compared with other models that build on the same data. These two findings collectively suggest highly localized spatial correlations at the beginning of the pandemic, yet this effect started to decline as the pandemic evolved. Figure 3 shows the geographic distribution of local coefficient estimates of GWR models for COVID-19 incidence rate associated with each explanatory variable. For each explanatory variable, we can observe a clear trend suggesting that the heterogeneity among coefficients became homogeneity throughout the time.

**Table 5.** Summary of the results from GWR model fitting

| July 15<sup>th</sup> |  |  |  |  |  |
| --- | --- | --- | --- | --- | --- |
| Covariates | Min. | Q1. | Median | Q3 | Max. |
| Male percentage | 6.73 | 7.68 | 8.11 | 8.50 | 8.98 |
| White percentage | -0.59 | -0.36 | -0.20 | -0.02 | 0.21 |
| Median age | -0.67 | -0.15 | 0.09 | 0.34 | 0.71 |
| College degree rate | 0.64 | 0.71 | 0.75 | 0.80 | 0.83 |
| Obesity rate | -0.81 | -0.49 | -0.32 | -0.10 | 0.22 |
| Unemployment rate | 1.82 | 2.15 | 2.36 | 2.66 | 3.18 |
| Uninsured rate | 0.48 | 0.76 | 0.93 | 1.20 | 1.58 |
| NP abundance | 0.17 | 0.20 | 0.21 | 0.23 | 0.24 |
| August 15<sup>th</sup> |  |  |  |  |  |
| Covariates | Min. | Q1. | Median | Q3 | Max. |
| Male percentage | 7.11 | 7.33 | 7.44 | 7.60 | 7.85 |
| White percentage | -0.67 | -0.65 | -0.63 | -0.61 | -0.59 |
| Median age | 0.32 | 0.41 | 0.44 | 0.50 | 0.57 |
| College degree rate | 0.48 | 0.49 | 0.49 | 0.50 | 0.50 |
| Obesity rate | -0.06 | -0.05 | -0.03 | -0.01 | 0.02 |
| Unemployment rate | 1.57 | 1.64 | 1.69 | 1.75 | 1.85 |
| Uninsured rate | -0.05 | -0.01 | 0.01 | 0.05 | 0.10 |
| NP abundance | 0.12 | 0.12 | 0.12 | 0.12 | 0.13 |
| September 15<sup>th</sup> |  |  |  |  |  |
| Covariates | Min. | Q1. | Median | Q3 | Max. |
| Male percentage | 9.28 | 9.44 | 9.55 | 9.69 | 9.90 |
| White percentage | -0.43 | -0.38 | -0.37 | -0.35 | -0.33 |
| Median age | 0.20 | 0.28 | 0.31 | 0.36 | 0.43 |
| College degree rate | 0.42 | 0.43 | 0.44 | 0.44 | 0.45 |
| Obesity rate | -0.07 | -0.05 | -0.04 | -0.02 | 0.01 |
| Unemployment rate | 1.77 | 1.83 | 1.87 | 1.93 | 2.02 |
| Uninsured rate | -0.22 | -0.17 | -0.14 | -0.11 | -0.05 |
| NP abundance | 0.12 | 0.12 | 0.13 | 0.13 | 0.13 |
| October 15<sup>th</sup> |  |  |  |  |  |
| Covariates | Min. | Q1. | Median | Q3 | Max. |
| Male percentage | 11.37 | 11.58 | 11.69 | 11.82 | 12.05 |
| White percentage | 0.07 | 0.11 | 0.13 | 0.15 | 0.17 |
| Median age | 0.20 | 0.24 | 0.27 | 0.29 | 0.35 |
| College degree rate | 0.30 | 0.31 | 0.32 | 0.33 | 0.33 |
| Obesity rate | 0.29 | 0.31 | 0.33 | 0.36 | 0.39 |
| Unemployment rate | 1.81 | 1.86 | 1.89 | 1.94 | 2.00 |
| Uninsured rate | -0.31 | -0.27 | -0.25 | -0.22 | -0.17 |
| NP abundance | 0.16 | 0.16 | 0.16 | 0.16 | 0.17 |
| November 15<sup>th</sup> |  |  |  |  |  |
| Covariates | Min. | Q1. | Median | Q3 | Max. |
| Male percentage | 11.96 | 12.15 | 12.24 | 12.37 | 12.57 |
| White percentage | 0.57 | 0.60 | 0.63 | 0.65 | 0.68 |
| Median age | 0.03 | 0.08 | 0.11 | 0.14 | 0.20 |
| College degree rate | 0.27 | 0.28 | 0.29 | 0.30 | 0.30 |
| Obesity rate | 0.53 | 0.55 | 0.57 | 0.59 | 0.64 |
| Unemployment rate | 1.91 | 1.95 | 1.98 | 2.03 | 2.09 |
| Uninsured rate | -0.51 | -0.47 | -0.45 | -0.43 | -0.39 |
| NP abundance | 0.16 | 0.17 | 0.17 | 0.17 | 0.17 |
| December 15<sup>th</sup> |  |  |  |  |  |
| Covariates | Min. | Q1. | Median | Q3 | Max. |
| Male percentage | 12.03 | 12.19 | 12.29 | 12.43 | 12.62 |
| White percentage | 1.14 | 1.18 | 1.21 | 1.23 | 1.27 |
| Median age | -0.18 | -0.13 | -0.10 | -0.07 | -0.02 |
| College degree rate | 0.28 | 0.29 | 0.30 | 0.31 | 0.32 |
| Obesity rate | 0.68 | 0.70 | 0.72 | 0.74 | 0.79 |
| Unemployment rate | 2.16 | 2.20 | 2.23 | 2.27 | 2.33 |
| Uninsured rate | -0.69 | -0.65 | -0.63 | -0.60 | -0.57 |
| NP abundance | 0.16 | 0.17 | 0.17 | 0.17 | 0.17 |

NP: nurse practitioner

**Table 6.** Summary of models’ AIC values across the time

|  | SEM | SLM | CAR Model | GWR Model |
| --- | --- | --- | --- | --- |
| **July 15** | 61.59 | 58.72 | 59.39 | 51.24 |
| **August 15** | 41.47 | 36.64 | 39.81 | 34.49 |
| **September 15** | 35.62 | 30.67 | 35.20 | 25.64 |
| **October 15** | 17.00 | 11.96 | 17.42 | 5.31 |
| **November 15** | 11.52 | 6.68 | 11.94 | -0.54 |
| **December 15** | 10.74 | 8.31 | 11.13 | -1.48 |

**Figure 3.**
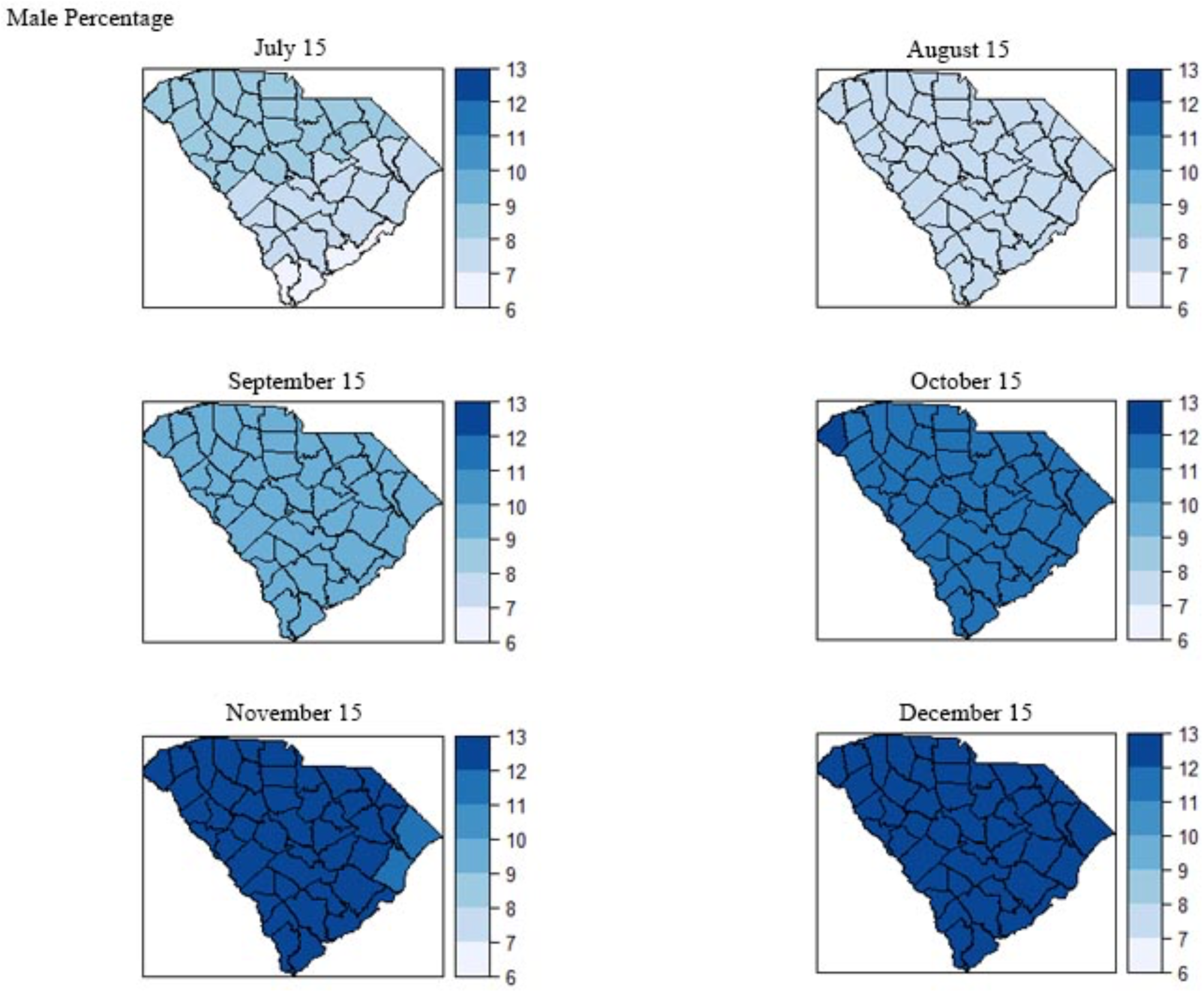
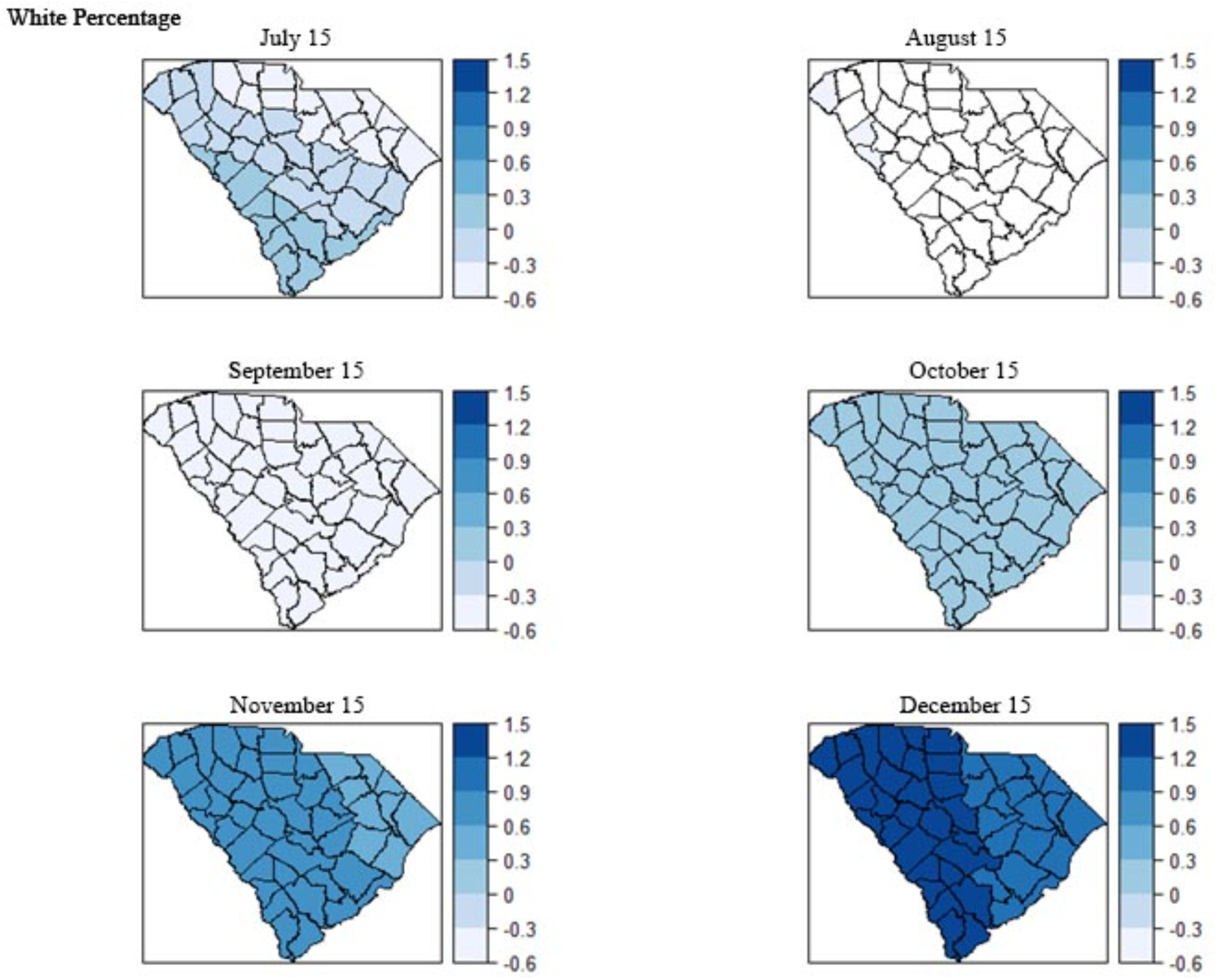
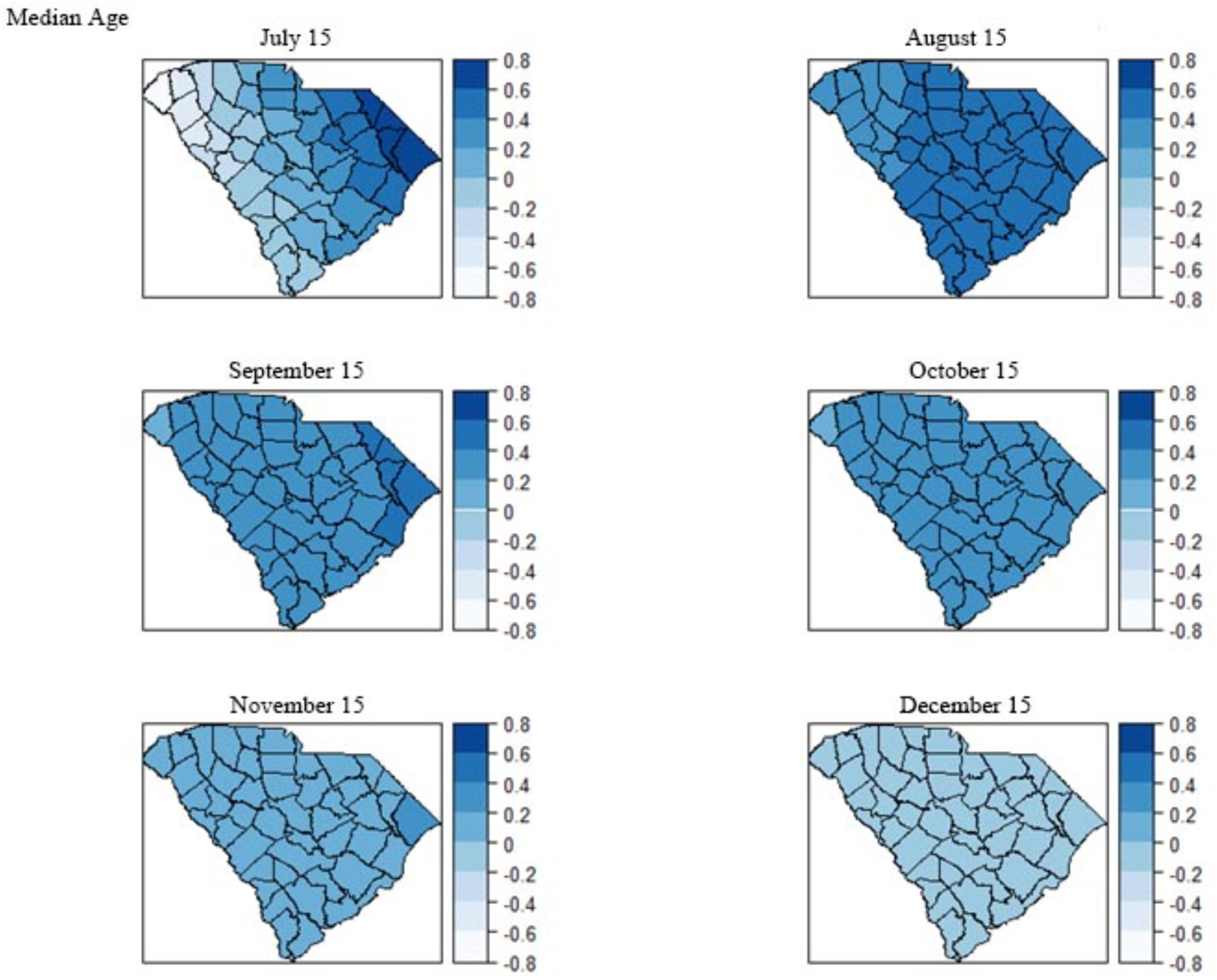
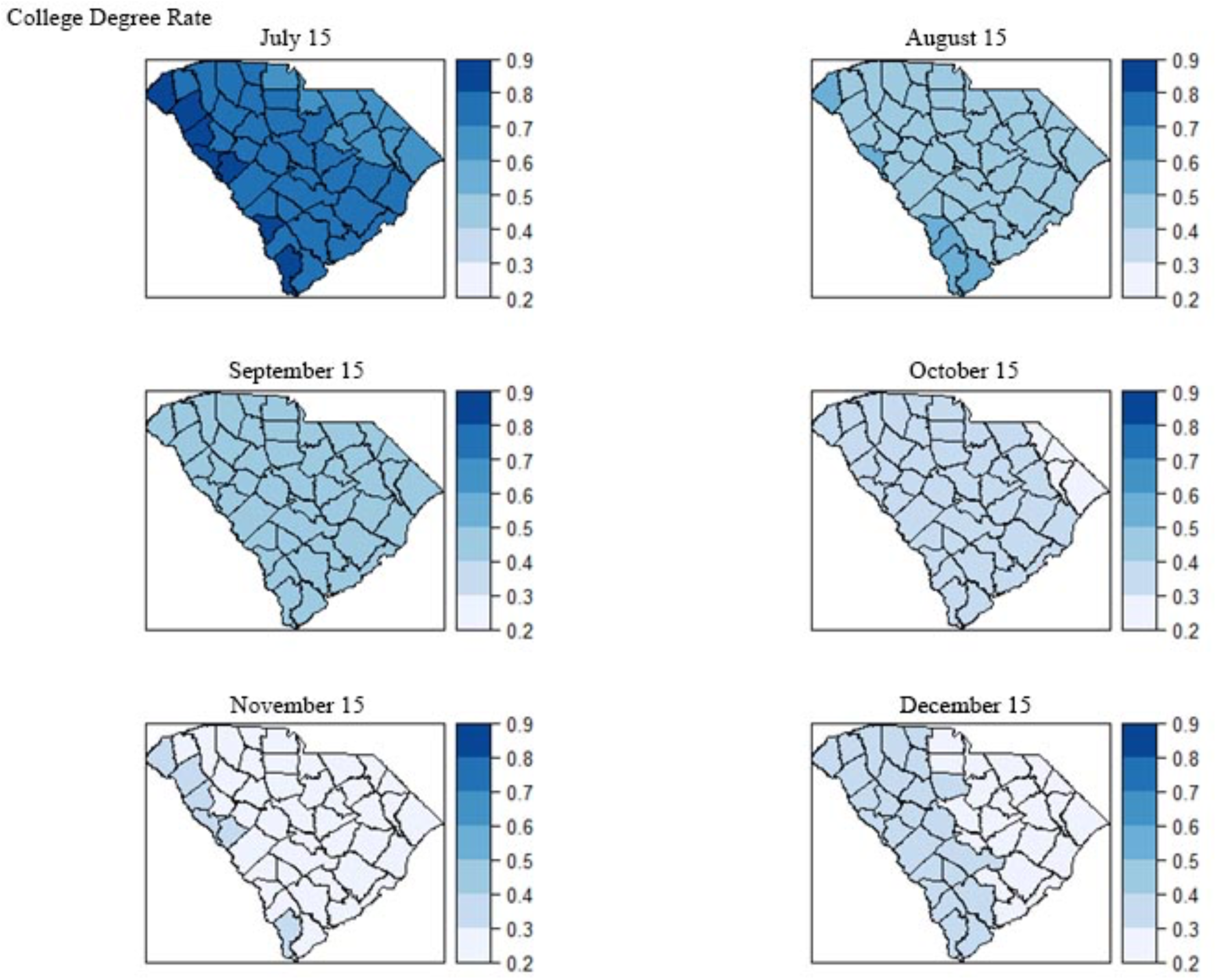
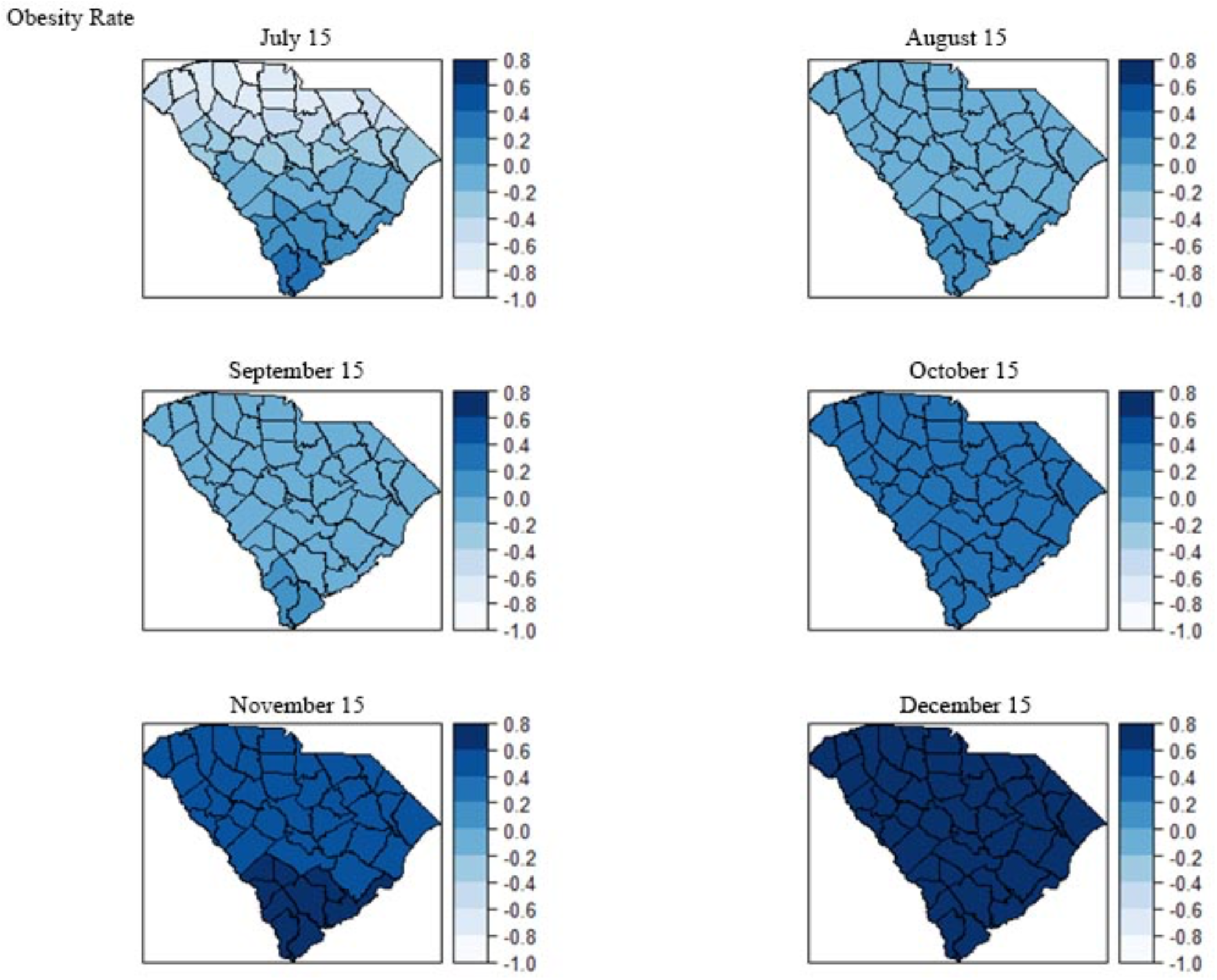
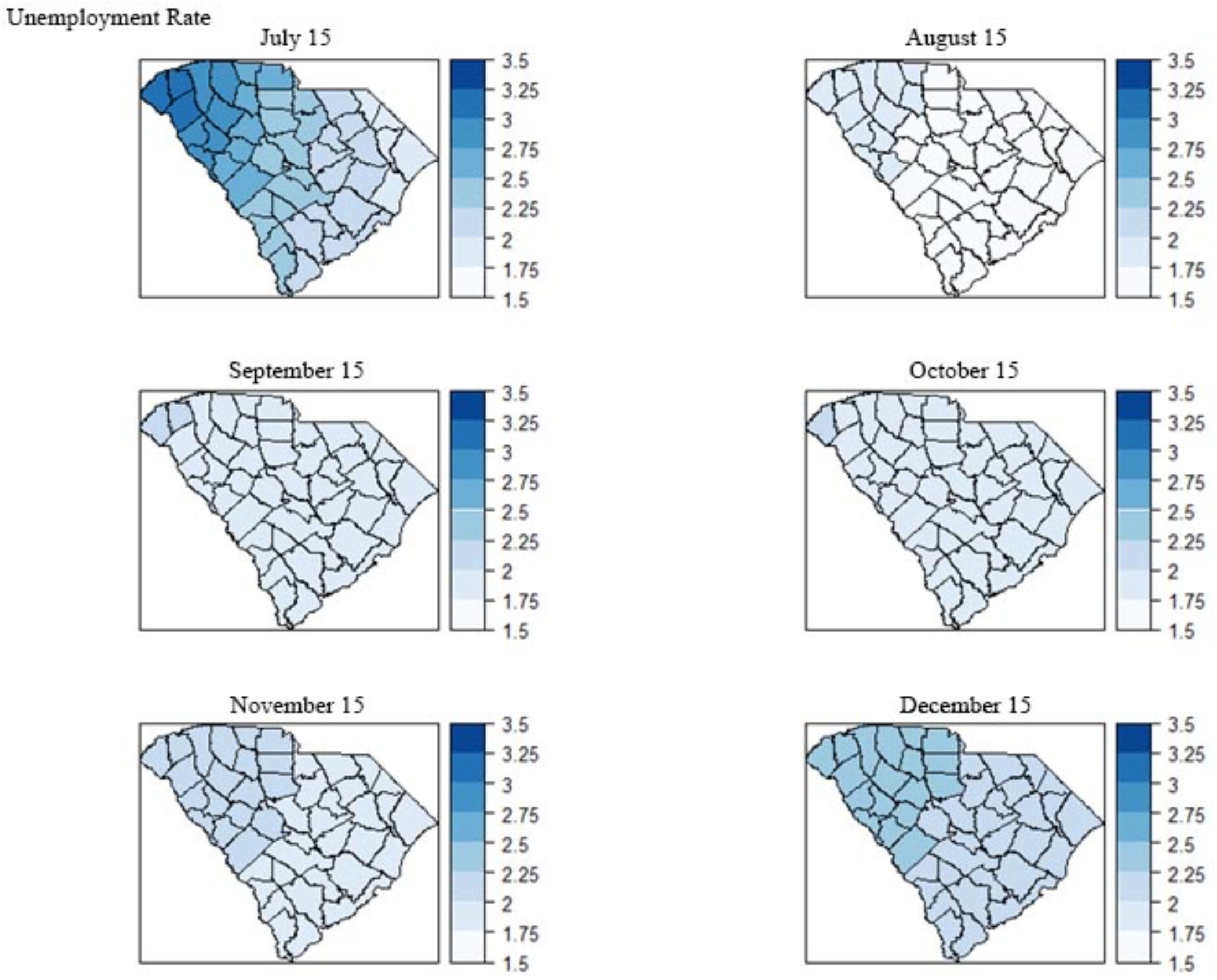
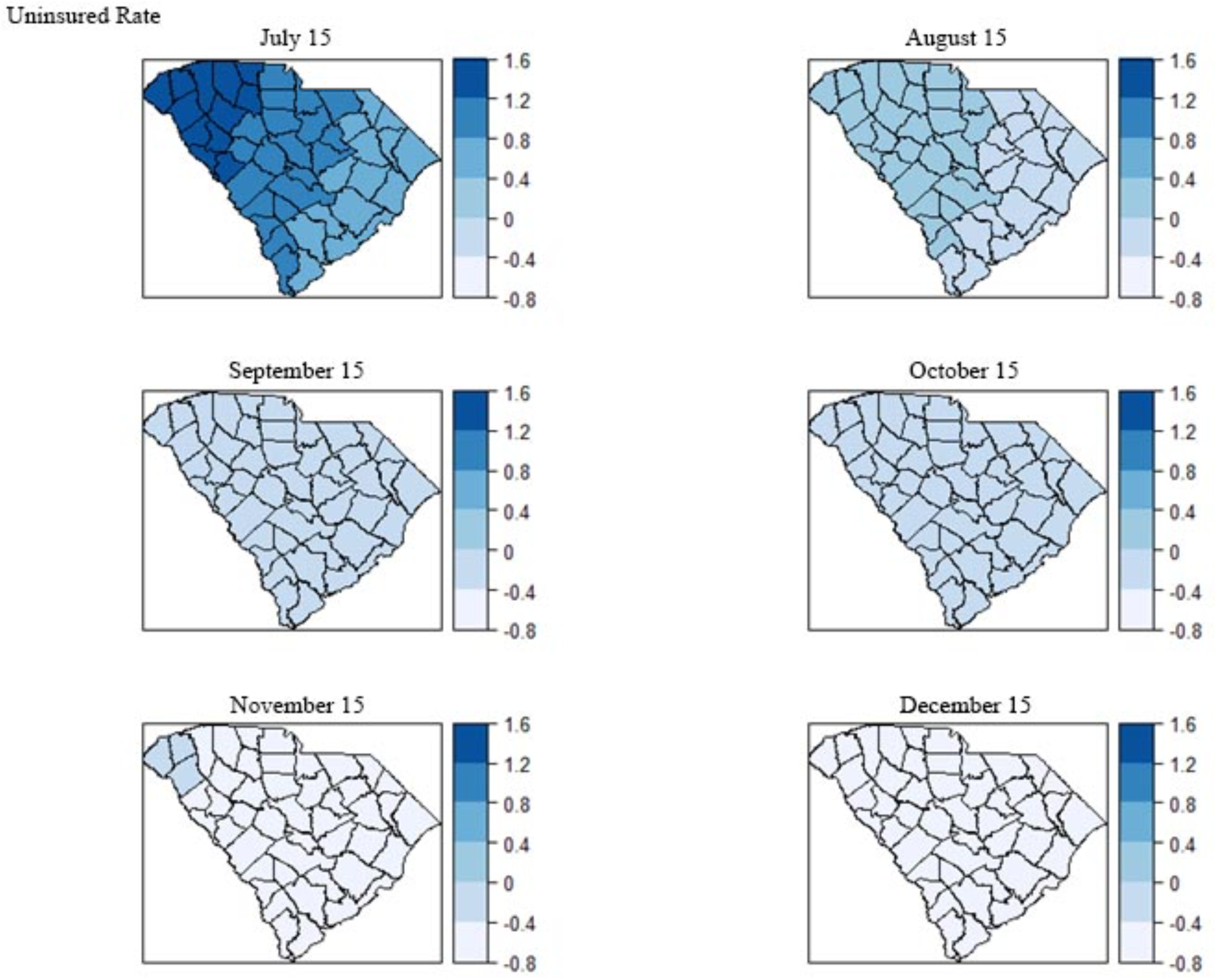
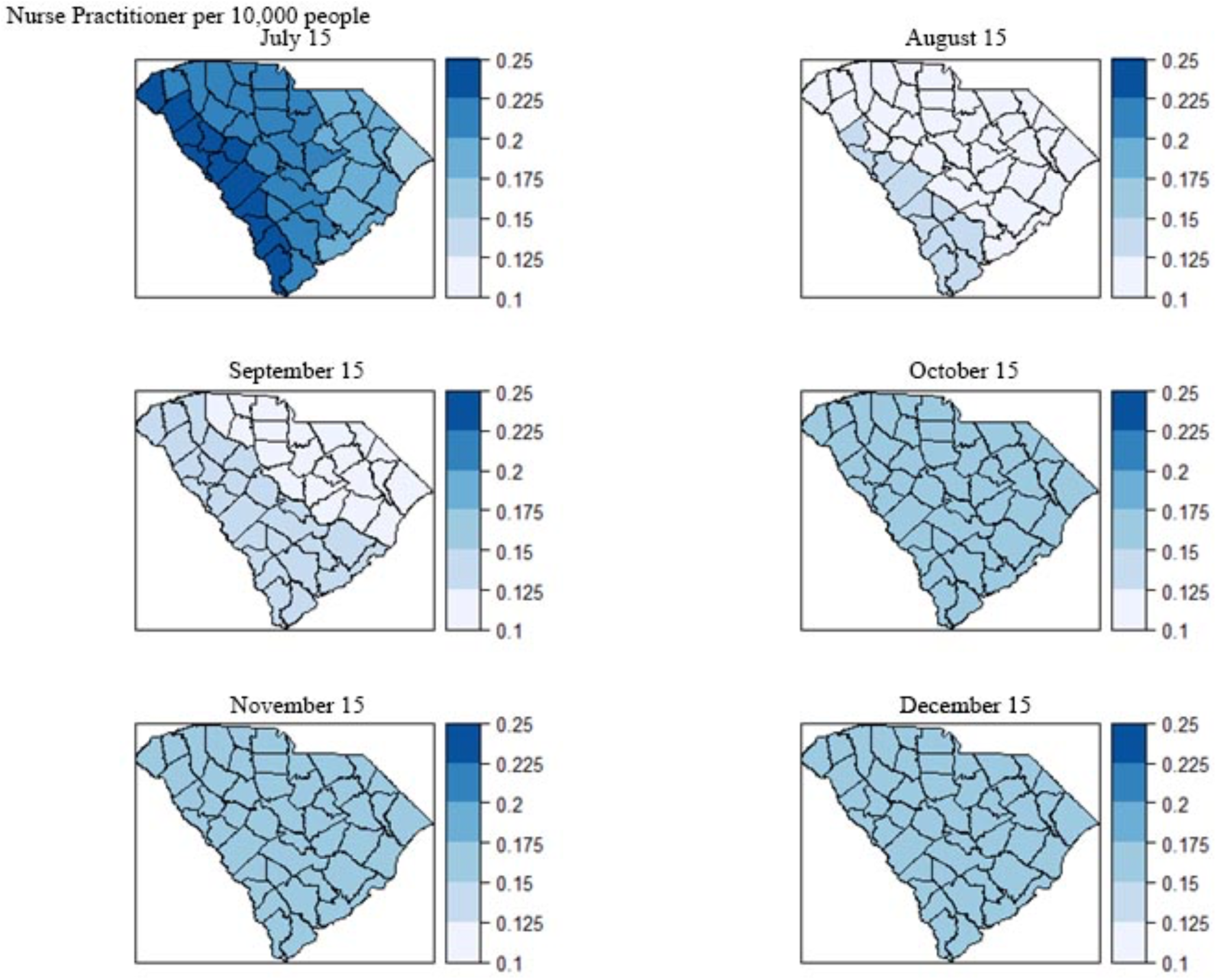
Geographic distribution of local coefficient estimates of GWR models for COVID-19 incidence rate associated with the explanatory variables

## Discussion

In this geospatial study, we adapted the socio-ecological vulnerability index from Snyder and Parks and compiled 15 variables within four categories of the index which could potentially explain the geographic patterns of COVID-19 transmission in SC.^36^ Our study resulted in three principal findings. First, our study validated that the spatial autocorrelations of COVID-19 incidence did exist at the county level in SC. The results from global models and local models are consistent with the initial observation of the distribution maps of covariates. Second, some PIDRs (e.g., male percentage, unemployment rate) have consistent spatial correlations with COVID-19 incidence over time while some other PIDRs (e.g., percentage of the white population, obesity rate) show divergent spatial correlations at different times of the pandemic, suggesting a critical role of the temporal dimension in the geospatial epidemiology of COVID-19 transmission. Third, the geospatial effect of PIDRs was strong at the beginning of the pandemic and started to decline as the infection cases continued to surge, suggesting the importance of early identification of critical PIDRs and intervention for the possible future outbreak of infectious diseases. Implications of these findings are discussed as follows.

Aligned with existing studies ^27,29^, two PIDRs (e.g., male percentage and unemployment rate) were found to be significantly associated with a higher risk of COVID-19 infection in global models including SEM, SLM, and CAR model. The higher risk of getting infected with COVID-19 among the male population can be explained by several gender-related factors.^39^ Genetically, males have a higher expression of angiotensin-converting enzyme-2 (ACE2), which could be the receptor for SARS-CoV-2.^40,41^ The immunological response of SAR-CoV-2 may be different between males and females.^42,43^ In addition, women were found to have a more responsible attitude of health behaviors towards COVID-19 than men.^18,44^ Higher unemployment rate reflects a higher socio-economic vulnerability of COVID-19 infection. People with the ability to work from home have been proven to be less likely infected with COVID-19 because of higher job security.^45,46^ On the contrary, unemployed people tend to be active in the labor market and are more likely to get infected.

We found that the white population was not statistically correlated with COVID-19 incidence from July to October and became positively correlated with COVID-19 incidence (all p < 0.01 for SEM, SLM, CAR) in December. To the best of our knowledge, this finding was not reported. We suspect that this finding was due to that the COVID-19 incidence rate is higher in large metropolitan areas (e.g., urban, suburban) in an early time of the pandemic (i.e., March-May 2020) and diffused to small and non-metropolitan areas in a later time where proportions of white are higher. ^30^ Among the 26 counties that are classified as metropolitan areas in South Carolina, only three of them have a white population of less than 50%, and five counties have a white population of less than 60%.^47,48^ Previous studies found that racial minorities had a higher risk of COVID-19 infection ^27,29,31^ but these findings have not been tested or interpreted by the temporal dimension of the pandemic. Cunningham and Wigfall reported that racial attitudes towards COVID-19 have a significant impact on the infection and mitigated the effect of racial difference, which also could explain our finding.^49^ In addition, our finding could be related to the finding that a higher proportion of white people took COVID-19 tests than other races in the latter months.^50^ Median age, college degree rate, obesity rate, uninsured rate, and NP abundance were not statistically correlated with COVID-19 infection rate.

Our findings suggested that early measures are critical to prevent the transmission of COVID-19 since the geographic differences in COVID-19 infection decreased over time, indicated by the decreasing AIC values across models longitudinally (Table 6). The decreasing AICs suggested a better fit of the model. The decrease in AICs of local model (i.e., GWR model) over time indicated the persistence of the non-stationary spatial autocorrelation. Although GWR models have lower AIC values compared with the global models, the coefficients of the variables in GWR models did not vary substantially, indicating small non-stationary effects. The small ranges of the coefficients geographically could be related to the insufficient granularity of the county-level data considering the study sample of South Carolina. Nevertheless, it is very interesting that the results from GWR models showed a pattern of the coefficients that the regional variances were decreasing during the study time frame. We used the same scale for each variable across the time to show the pattern more clearly.

This study is among the first to examine geospatial patterns on COVID-19 infection as well as the corresponding PIDRs. Most studies have been focusing on patients with different levels of severity with COVID-19, which limit the opportunities of examing possible disparities and PIDRs in COVID-19 infection.^51^ For example, older adults, people with certain medical conditions, and pregnant women were found to be associated with a higher risk of severe illnesses of COVID-19, while our study found that the male population and unemployment rate were at higher risk of COVID-19 infection.^51^ Intuitively, the PIDR set for severe illnesses of COVID-19 is related to the physical condition of the patients and the PIDR set for COVID-19 infection is influenced by the demographic and socioeconomic factors. Compared with PIDRs for severe illnesses, the PIDRs for infection are highly sensitive to geographic regions and temporal dynamics of the pandemic because the transmission of COVID-19 depends on the activity of people. PIDRs for COVID-19 infection provide important information for developing interventions on targeted populations who share the same PIDRs at the beginning of the pandemic, so as to contain the early-stage transmission and potential consequences of population health in future infectious disease outbreaks.

Our study suffered from a few limitations. First, we did not use longitudinal measures of PIDRs due to limited public health surveillance. Second, data for COVID-19 testing rates for each race are not available when examining the racial differences. Third, due to the limited data access, we used county-level data rather in this study whereas using zip code-level data would have offered a better granularity of data in the statistical models. At last, variables used in this study may not be complete because we primarily adopted the framework from Snyder and Parks to conceptualize the study where the framework was designed for studying COVID-19 infection. Future studies could also integrate other frameworks with a better coverage of Social Vulnerability Index (SVI), a varible for exploring the negative effects in communities towards hazardous events.^52^

## Conclusion

Our study found that the geospatial distribution of COVID-19 incidence was constantly influenced by several key PIDRs including male percentage and unemployment. PIRDs such as white percentage and obesity rate were negatively correlated with COVID-19 incidence at the beginning of the pandemic and were then became positively correlated with COVID-19 incidence. These identified PIDRs are different risk factors as compared to risk factors found to be associated with poor clinical outcomes (e.g., severity, mortality) of patients who are connected with medical care. Our findings support found disparities in COVID-19 transmission and suggest newly identified temporal dynamics in specific PIDRs such as white percentage and obesity rate. These findings are subject to biases caused by limited data access and should be considered provisional guides to the temporal geospatial epidemiology of COVID-19 transmission and underlying PIDRs of the pandemic in South Carolina.

## Data Availability

All data referred to in this manuscript are publicly available.

